# The association between asthma and the risk of macular degeneration: findings from the English longitudinal study of ageing

**DOI:** 10.64898/2026.03.12.26348266

**Authors:** Yini Yang, Jingwei Li

## Abstract

In recent years, researchers have paid increasing attention on potential associations between respiratory and ocular diseases. To examine whether asthma is independently associated with macular degeneration (MD) and whether asthma can serve as a predictor of MD using data from the English Longitudinal Study of Ageing (ELSA). Data from the 2020-2021 wave of ELSA were analyzed. Statistical tests were performed on participants’ baseline characteristics. Multivariable logistic regression, stratified analysis, ROC curve analysis, smoothing curves and sensitivity analysis were conducted to assess the association, stability, predictive performance, dose-response relationship and robustness. A total of 6,703 participants were included. After adjusting for covariates, age and asthma were significantly associated with MD (p < 0.05), while diet and sex were not. Asthma was consistently linked to increased MD risk across three regression models (OR > 1, p < 0.05), with the association persisting in stratified analyses. ROC analysis showed moderate predictive performance (AUC = 0.757), and a positive dose-response relationship was observed. Sensitivity analyses confirmed the robustness of the association. Asthma may independently increase MD risk, providing novel insights into their relationship and implications for clinical risk stratification and preventive strategies.

## 1. Introduction

Macular degeneration (MD) is a chronic degenerative retinal disease characterized by progressive degeneration of retinal pigment epithelium (RPE) cells and photoreceptors, leading to irreversible central vision loss and representing a major cause of blindness globally[1, 2]. Clinicians always categorize MD into two main forms: age-related macular degeneration (AMD) and inherited retinal degeneration (IRD). AMD represents over 90% of MD cases and affects 12%-17% of individuals aged 50 years or older worldwide[3–5]. Due to global aging trends, AMD prevalence is expected to rise by 30% by 2050, placing great pressure on healthcare systems[5]. The pathogenesis of AMD involves oxidative stress, choroidal neovascularization, and inflammation. Key risk factors include aging, smoking, obesity, hypertension, cardiovascular disease, and genetic predisposition[6–8]. Current treatment mainly targets on wet AMD, the more rapidly progressive and vision-threatening subtype, with intravitreal anti-VEGF therapy[9, 10]. However, this approach requires frequent injections and shows limited effectiveness for dry AMD[11, 12]. The 2024 American Academy of Ophthalmology (AAO) guidelines emphasize the clinical value of early screening and preventive measures for high-risk groups to reduce AMD incidence and preserve vision[13].

Asthma is a common, heterogeneous airway disease marked by chronic inflammation and increased airway hyperresponsiveness. The main clinical symptoms of asthma are recurrent wheezing, chest tightness, and shortness of breath[14–16]. Allergic asthma is the most common subtype, accounting for approximately 50%-70% of all cases[17, 18]. Recent studies has explored a potential association between allergic diseases (including asthma) and MD. A study using Taiwan’s National Health Insurance Database reported that people with prior allergic diseases, such as asthma, had a 32% higher risk of developing AMD (odds ratio [OR]=1.32, 95% confidence intervals [CI] 1.18-1.48) [19]. Similarly, a large-scale prospective Korean cohort study revealed that AMD patients had a 25% higher risk of developing allergic diseases, including asthma (hazard ratio [HR]=1.25, 95% CI 1.12-1.39)[20]. These findings suggest a potential bidirectional association between asthma and MD, though the strength of this association warrants further investigation.

The English Longitudinal Study of Ageing (ELSA) is a comprehensive long-term study that monitors health, social, and psychological changes in adults aged over 50 in the United Kingdom[21]. Previous research utilizing ELSA dataset has already identified associations between visual impairment and adverse health outcomes, such as cognitive decline or physical inactivity[22, 23]. However, the specific association between asthma and MD has not been systematically examined in the UK. To address this gap, we analyzed data from the 2020-2021 ELSA wave to assess this association through multivariable logistic regression. We further evaluated the predictive value of this association through ROC curve analysis, risk stratification, and sensitivity analyses. Our objective was to investigate the association between asthma and MD in British adults aged 50 years and older. We also aimed to evaluate whether asthma status could inform targeted early screening or prevention strategies for MD.

## 2. Materials and methods

### 2.1 Study population inclusion

The ELSA data used in this study were obtained from the 2020-2021 wave of ELSA. All related data and materials are publicly available through the official website: https://www.elsa-project.ac.uk. All non-sensitive data can be requested via the UK Data Service platform (https://beta.ukdataservice.ac.uk/datacatalogue/series/series?id<u>= 200011</u>). The study strictly adhered to ethical approval from the UK National Health Service Research Ethics Committee, and all participants provided written informed consent. All procedures followed relevant ethical guidelines and data security standards to ensure data authenticity, integrity, and participant confidentiality. Exclusion criteria included: (1) Participants aged <50 years (the lower age limit for ELSA enrollment); (2) Individuals with missing information on asthma status; (3) Those who did not meet the criteria for macular degeneration or with missing related information; (4) Participants with missing or refused data on key covariates. The inclusion and exclusion process is illustrated in **Fig 1**. After screening, 6,703 eligible respondents were included in subsequent analysis.

**Fig 1.**
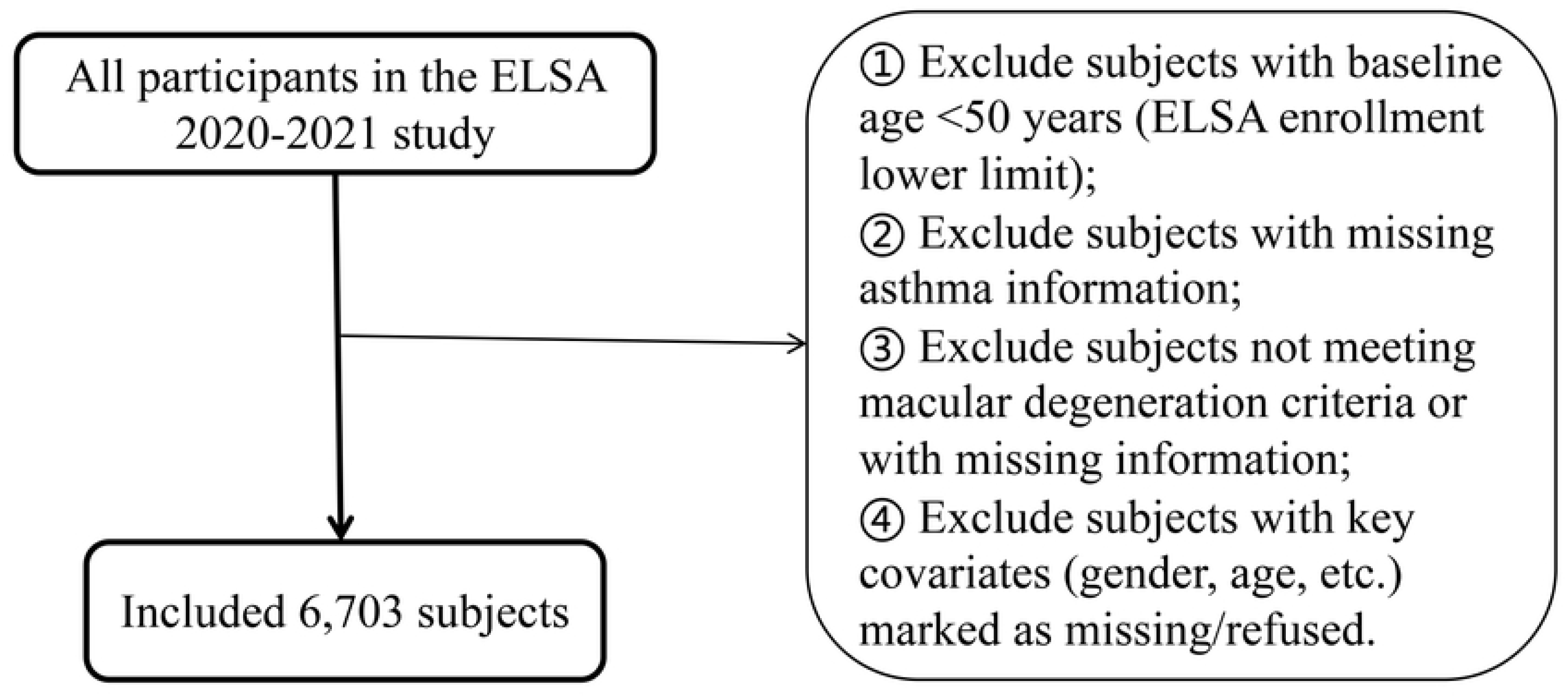
Flowchart of participant inclusion and exclusion.

### 2.2 Definitions of exposure, outcome, and covariates

In the ELSA database, the exposure variable was defined based on physician diagnoses and self-reported responses to the question “Chronic: Diagnosed with asthma” (code: HEEVERAS), categorized as: 1. YES (included in analysis), 2. NO (excluded from analysis). The outcome variable was determined based on physician diagnoses and self-reported responses to the question “Ever been told has macular degeneration by a doctor?” (code: HEEVERMD), categorized as: 1. YES, 2. NO. Covariates included age, sex, race, marital status, smoking status, diet, hypertension, physical activities, health status, and diabetes. Detailed descriptions of these covariates are provided in Supplementary **S1 Table**.

### 2.3 Statistical analysis

Data from the 2020-2021 wave of ELSA were analyzed to compare baseline characteristics between participants with and without MD. Categorical baseline characteristics were described using frequencies (n) and percentages (%). Continuous variables were summarized as means ± standard deviations when normally distributed, or medians (interquartile ranges) when distributions were skewed. Baseline comparisons were performed using the “tableone” package (v0.13.2)[24]. Two-sample t tests were applied to continuous variables, while chi-square tests were used for categorical variables to assess differences in potential confounding factors between groups. These descriptive comparisons were used to characterize baseline differences between groups before association analyses.

Associations between asthma and MD risk were evaluated using progressively adjusted logistic regression models implemented in the “stats” package (v 4.3.3)[25]. Three logistic regression models were specified. Model 1 was a unadjusted baseline model, including only the exposure variable (asthma) and the outcome variable (MD). Model 2 extended Model 1 by adjusting for basic demographic factors, including sex, age, and ethnicity. Model 3 further adjusted for all covariates on the basis of Model 2 variables. Associations between variables and MD risk were quantified using ORs with 95% CIs. OR > 1 indicated an increased risk of MD, OR < 1 suggested a potential protective association, OR = 1 indicated no association.

To further examine whether the association between asthma and MD was consistent across different population subgroups, subgroup analyses were performed based on Model 3 using the “jstable” package (v 1.3.13)[26]. Forest plots were generated using the “forestploter” package (version 1.1.3) to visualize the association strength (OR values) and 95% CIs between the exposure factor (asthma) and outcome (MD) across different subgroups[27]. The predictive performance of asthma for MD was evaluated using ROC curve analysis based on Model 3. ROC curves were plotted with the “pROC” package (v 1.18.5)[28], and the AUC was calculated as a summary measure of discrimination. To explore a potential dose-response relationship between asthma and MD, weighted logistic regression analyses accounting for complex survey was performed based on Model 3 using the “survey” package (v 4.4.2)[29]. Smoothed curves were plotted using the “ggplot2” package (v 3.3.5) to illustrate the shape of this association[30]. Sensitivity analyses were conducted to assess the robustness of the association between asthma and MD. The analyses were performed using the “sensemakr” package (v 0.1.6) based on Model 3[31]. In addition, univariate benchmark analyses were applied to contextualize the potential impact of unobserved confounders.

All statistical analyses were performed using R software (version 4.2.2). A two-sided p value < 0.05 was considered statistically significant.

## Results

### 3.1 Baseline characteristics of the participants

The analysis included 6,703 participants from the 2020–2021 wave of the ELSA study. Of these, 314 were classified into the MD group and 6,389 into the non-MD group. Baseline characteristic analysis indicated significant differences between participants with and without MD for asthma (p < 0.05) as well as all other covariates, except diet and sex. **(Table 1)**.

**Table 1.** Baseline characteristics of participants stratified by macular degeneration (MD) status: Columns 1 and 2 list the variables and their respective categories/levels. Columns 3 and 4 display the sample size (n) and percentage of the control and MD group at each level. Column 5 presents the P-value for between-group differences. Categorical variables are expressed as percentages, while continuous variables are presented as mean ± standard deviation (SD).

|  | level | control | disease | p |
| --- | --- | --- | --- | --- |
|  | n | 6389 | 314 |  |
| asthma (%) | No | 5421 (84.8) | 250 (79.6) | 0.015 |
|  | Yes | 968 (15.2) | 64 (20.4) |  |
| high_blood_pressure (%) | No | 3989 (62.4) | 152 (48.4) | <0.001 |
|  | Yes | 2400 (37.6) | 162 (51.6) |  |
| diabetes (%) | No | 5681 (88.9) | 247 (78.7) | <0.001 |
|  | Yes | 708 (11.1) | 67 (21.3) |  |
| Age (%) | <=70 | 3944 (61.7) | 77 (24.5) | <0.001 |
|  | >70 | 2445 (38.3) | 237 (75.5) |  |
| Sex (%) | Female | 3561 (55.7) | 188 (59.9) | 0.167 |
|  | Male | 2828 (44.3) | 126 (40.1) |  |
| Race (%) | Non-White | 379 ( 5.9) | 6 ( 1.9) | 0.004 |
|  | White | 6010 (94.1) | 308 (98.1) |  |
| Married (%) | Divorced | 771 (12.1) | 35 (11.1) | <0.001 |
|  | Married | 3512 (55.0) | 171 (54.5) |  |
|  | Remarried | 736 (11.5) | 28 ( 8.9) |  |
|  | Separated | 100 ( 1.6) | 3 ( 1.0) |  |
|  | Single | 616 ( 9.6) | 18 ( 5.7) |  |
|  | Widowed | 654 (10.2) | 59 (18.8) |  |
| Diet (mean (SD)) |  | 94.68 (69.74) | 86.99 (64.50) | 0.056 |
| Smoking (%) | No | 2745 (43.0) | 128 (40.8) | 0.041 |
|  | Unknown | 3326 (52.1) | 179 (57.0) |  |
|  | Yes | 318 ( 5.0) | 7 ( 2.2) |  |
| Health (%) | Bad | 1145 (17.9) | 87 (27.7) | <0.001 |
|  | Fair | 2071 (32.4) | 103 (32.8) |  |
|  | Good | 1935 (30.3) | 75 (23.9) |  |
|  | Very bad | 502 ( 7.9) | 38 (12.1) |  |
|  | Very good | 736 (11.5) | 11 ( 3.5) |  |
| Activities (%) | Hardly ever, or never | 3853 (60.3) | 238 (75.8) | <0.001 |
|  | More than once a week | 1468 (23.0) | 41 (13.1) |  |
|  | Once a week | 592 ( 9.3) | 14 ( 4.5) |  |
|  | One to three times a month | 476 ( 7.5) | 21 ( 6.7) |  |

### 3.2 Risk association analysis between MD and asthma

The association between asthma and MD was evaluated using three logistic regression models. In the unadjusted Model 1, asthma was associated with increased odds of MD (OR = 1.42, 95% CI: 1.079-1.868, p < 0.05). After adjustment for sex, age, and ethnicity in Model 2, the association remained significant (OR = 1.457, 95% CI: 1.107-1.919, p < 0.05). After adjustment for all remaining covariates (smoking status, diet, physical activity, health status, hypertension, diabetes) in Model 3, asthma remained significantly associated with a higher risk of MD (OR = 1.389, 95% CI: 1.054-1.831, p < 0.05) **(Table 2)**.

**Table 2.** Association between asthma and MD: Column 1 lists the exposure factor (asthma). Column 2 shows the three multivariable logistic regression models. Column 3 presents the Odds Ratio (OR), indicating the strength of the association between asthma and MD risk. Column 4 displays the corresponding 95% Confidence Interval (CI) for the OR. Column 5 shows the P-value indicating the statistical significance of the association between the exposure factor (asthma) and outcome event (MD).

|  |  | OR | 95% CI | *p*_value |
| --- | --- | --- | --- | --- |
| asthma | model 1 | 1.42 | (1.079-1.868) | 0.01234 |
|  | model 2 | 1.457 | (1.107-1.919) | 0.007286 |
|  | model 3 | 1.389 | (1.054-1.831) | 0.01962 |

Across Models 1-3, asthma showed a consistent positive association with increased MD risk, with effect estimates remaining similar after adjustment for demographic, lifestyle, and health status covariates. Overall, asthma was positively associated with MD risk after adjustment for multiple covariates.

### 3.3 Stratified analysis of the association between asthma and MD risk

Stratified logistic regression analyses based on Model 3 were performed to assess potential heterogeneity of the association between asthma and MD across different subgroups. Across subgroups including age, sex, ethnicity, smoking status, diet, and hypertension, ORs were generally greater than 1, with substantial overlap in 95% CIs **(Fig 2)**. In participants aged over 70 years, the association appeared stronger (OR = 1.57, 95% CI: 1.12–2.19, p < 0.05) compared with the overall sample. Taken together, in most subgroups, asthma showed a positive association between MD, although the magnitude of the association varied across different demographic and clinical characteristics.

**Fig 2.**
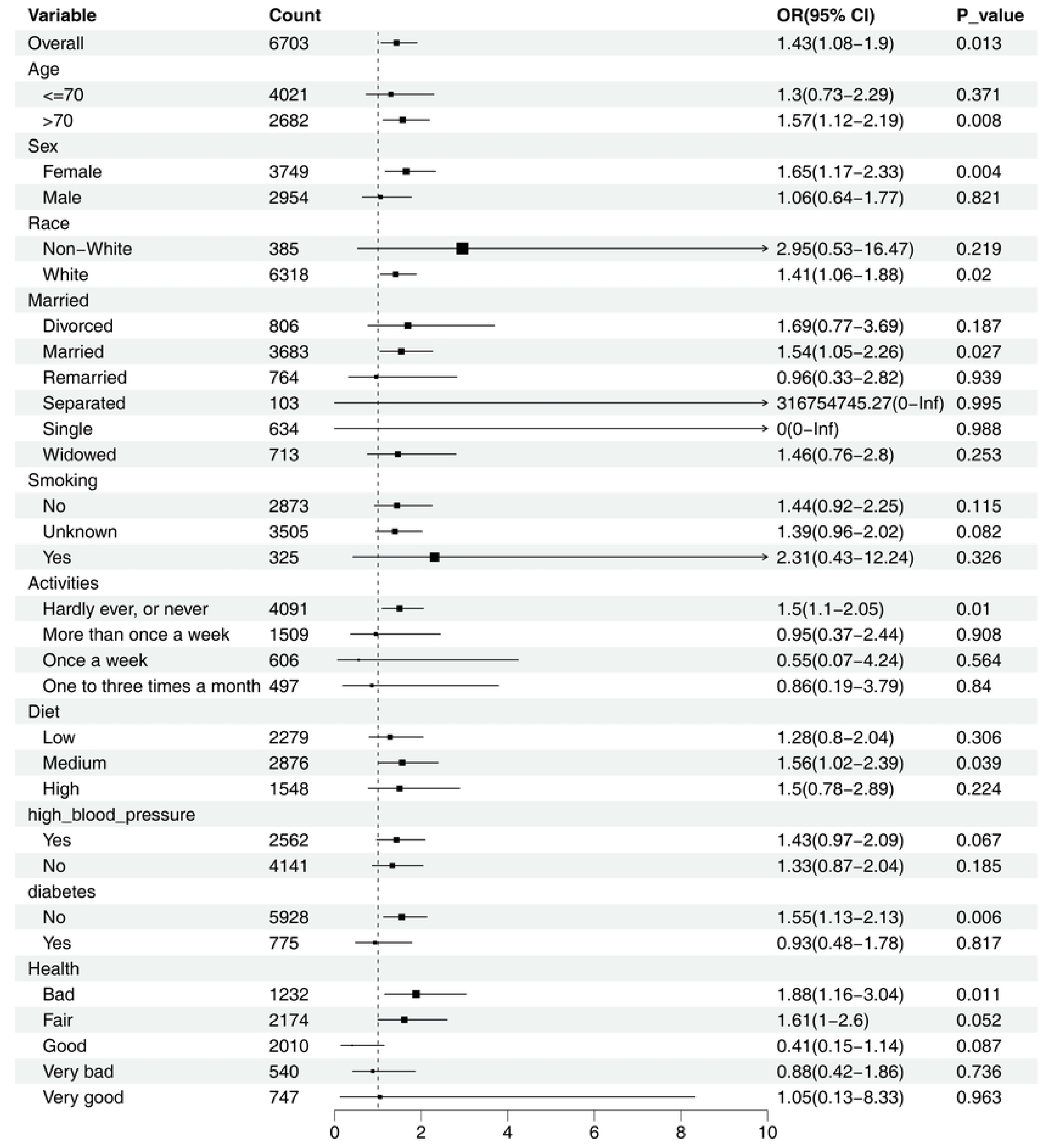
Stratified analysis of the association between asthma and macular degeneration (MD) risk: The figure displays the odds ratios (ORs) and relative 95% confidence intervals (CIs) for the association between exposure (asthma) and outcome (MD) across different subgroups. The vertical dashed line indicates the reference line at OR=1, indicating no association. Each square represents the OR of a specific subgroup, with larger squares reflecting higher OR values. Horizontal lines correspond to 95% CIs. If a 95% CI does not cross the reference line (OR = 1), the association between exposure (asthma) and outcome (MD) is considered statistically significant (p < 0.05) in that subgroup.

### 3.4 ROC curve evaluation of predictive performance

ROC curve analysis based on Model 3 was constructed to evaluate the predictive efficacy of asthma for MD. The AUC was 0.757 **(Fig 3)**. The result described the discriminatory performance of asthma for MD within the fully adjusted model.

**Fig 3.**
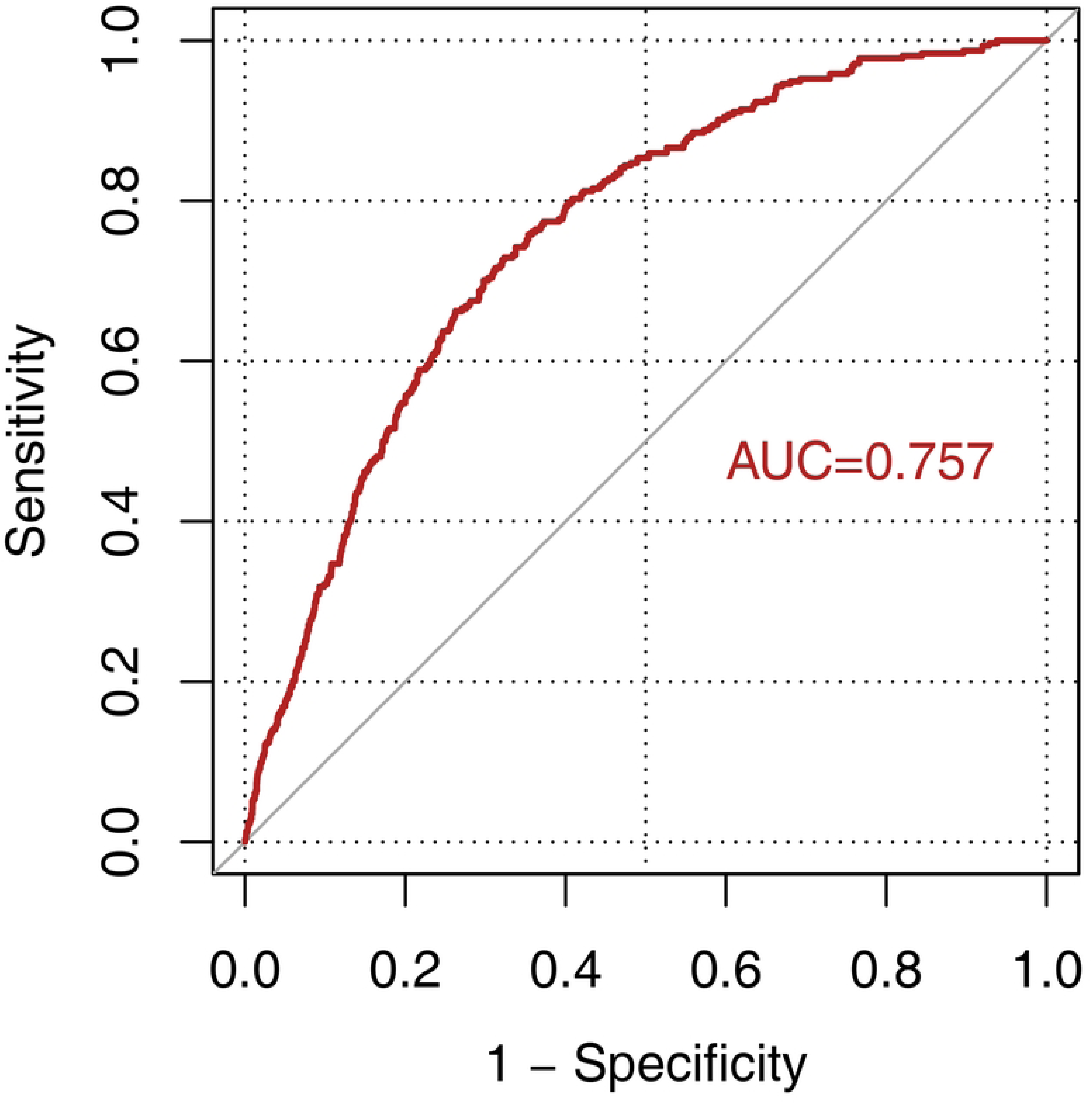
Receiver operating characteristic (ROC) analysis curve for the prediction of MD: The x-axis represents 1-specificity (false positive rate), and the y-axis represents sensitivity (true positive rate). The red curve illustrates the discriminative performance of the model across different thresholds, while the diagonal line serves as the reference (representing random classification).

### 3.5 Smooth curve fitting

A smooth curve analysis based on Model 3 was fitted to explore the relationship between asthma and MD. The smoothed curve showed a positive association between asthma and MD across the observed range of values **(Fig 4)**. No clear deviation from linearity was observed.

**Fig 4.**
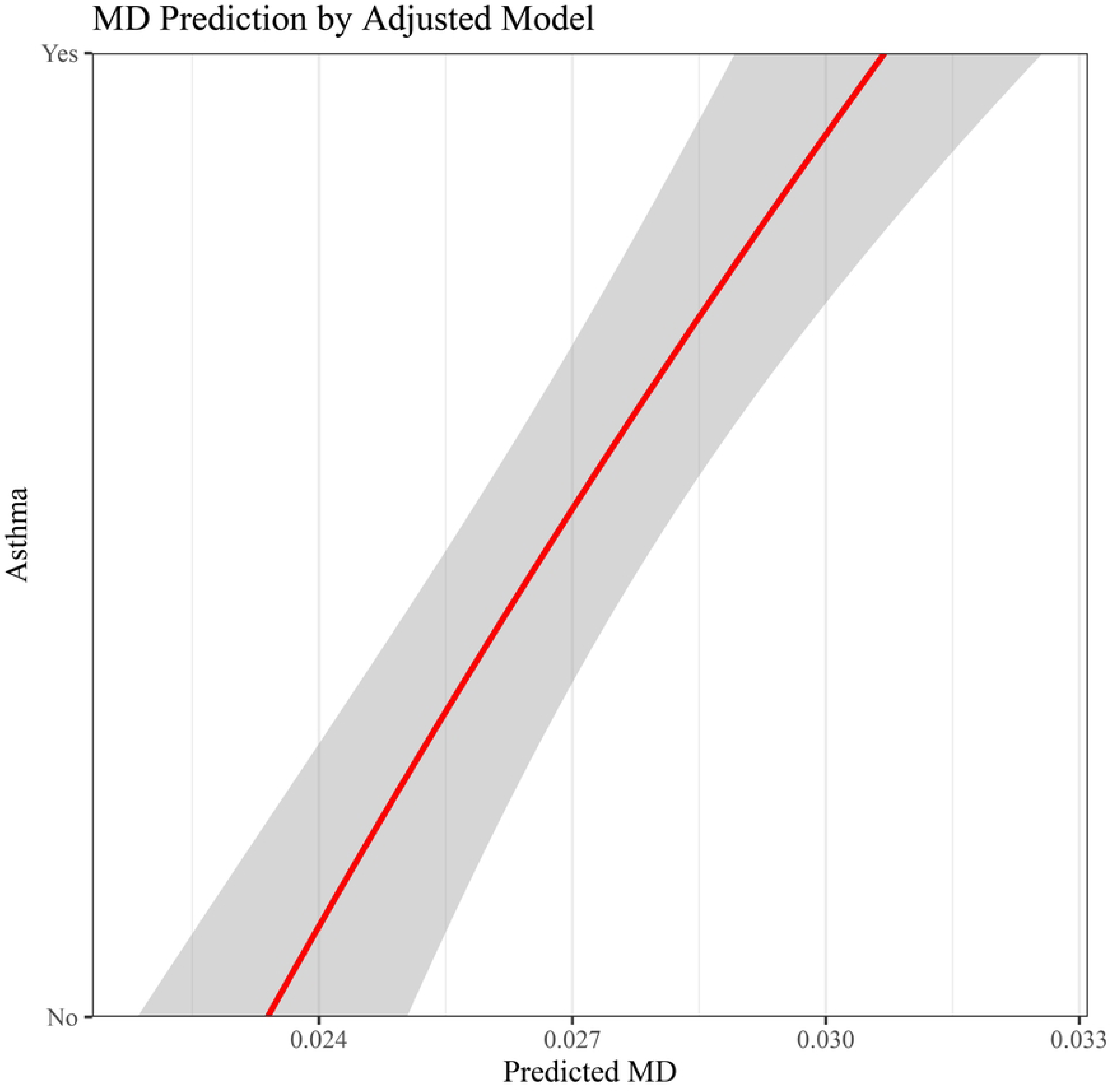
Smooth curve fitting for dose-response relationship between asthma and MD: The x-axis represents the predicted probability of MD, and the y-axis represents asthma status (No: without asthma, Yes: with asthma).

### 3.6 Sensitivity analysis

Sensitivity analyses based on Model 3 showed a positive association between asthma and MD after full adjustment for covariates (β = 0.017, t = 2.338). Sensitivity metrics suggested that the observed association could be influenced by unmeasured confounding. The robustness value (RVq = 1) was 2.8%, indicating that unobserved confounders would need to explain more than 2.8% of the residual variance in both the exposure (asthma) and outcome (MD) to fully attenuate the estimated association. The robustness value for maintaining statistical significance (RVq = 1, α = 0.05) was 0.5%, suggesting that an unobserved confounder would need to explain more than 0.5% of the residual variance to render the association statistically non-significant **(Table 3)**. Univariate benchmark analyses showed heterogeneity in sensitivity across different benchmark covariates. Benchmark related to age showed the largest influence. An unobserved confounder with three times the strength of age could reduce the regression coefficient β from 0.0129 to 0.0058, attenuating the effect size while maintaining statistical significance. Benchmarks related to sex, health status, and physical activity also exerted a notable influence, while those for smoking status, marital status, and diet showed minimal impact **(Table 4)**. In summary, sensitivity analyses indicated that the estimated association between asthma and MD may be influenced by unmeasured confoundings, particularly variables related to age, sex, and health status.

**Table 3.** Sensitivity statistics for the association between asthma and MD: Treatment: name of the exposure variable (asthma). Est.: original coefficient estimate for the effect of asthma on MD in the fully adjusted main model. S.E.: standard error. t-value: t-statistic value. R²_{Y∼D|X}: partial coefficient of determination of the treatment variable on the outcome, after controlling for observed variables (0.00%, as all relevant covariates were already included in the model). RVq = 1: robustness value, indicating the strength of unobserved confounding required to reduce the point estimate to 0. RVq = 1, α = 0.05: robustness value for statistical significance, indicating the strength of unobserved confounding required to render the estimate statistically non-significant. Note: Bias bounds derived from demographic benchmarks provide specific proportions of residual variance that unobserved confounders would need to explain simultaneously in both the treatment and the outcome.

| Table 3 Sensitivity statistics |  |  |  |  |  |  |
| --- | --- | --- | --- | --- | --- | --- |
| Outcome:MD |  |  |  |  |  |  |
| Treatment | Est. | S.E. | t-value | $R^2_{Y \sim D\vert X}$ | $RV_q = 1$ | $RV_q = 1, \alpha = 0.05$ |
| *asthma* | 0.017 | 0.007 | 2.338 | 0.10% | 2.80% | 0.50% |
| Note: df = 6691; Bound (1x Age): $R^2_{Y \sim Z\vert X,D} = 2.5\%$, $R^2_{D \sim Z\vert X} = 0.2\%$ | | | | | | |

**Table 4.**
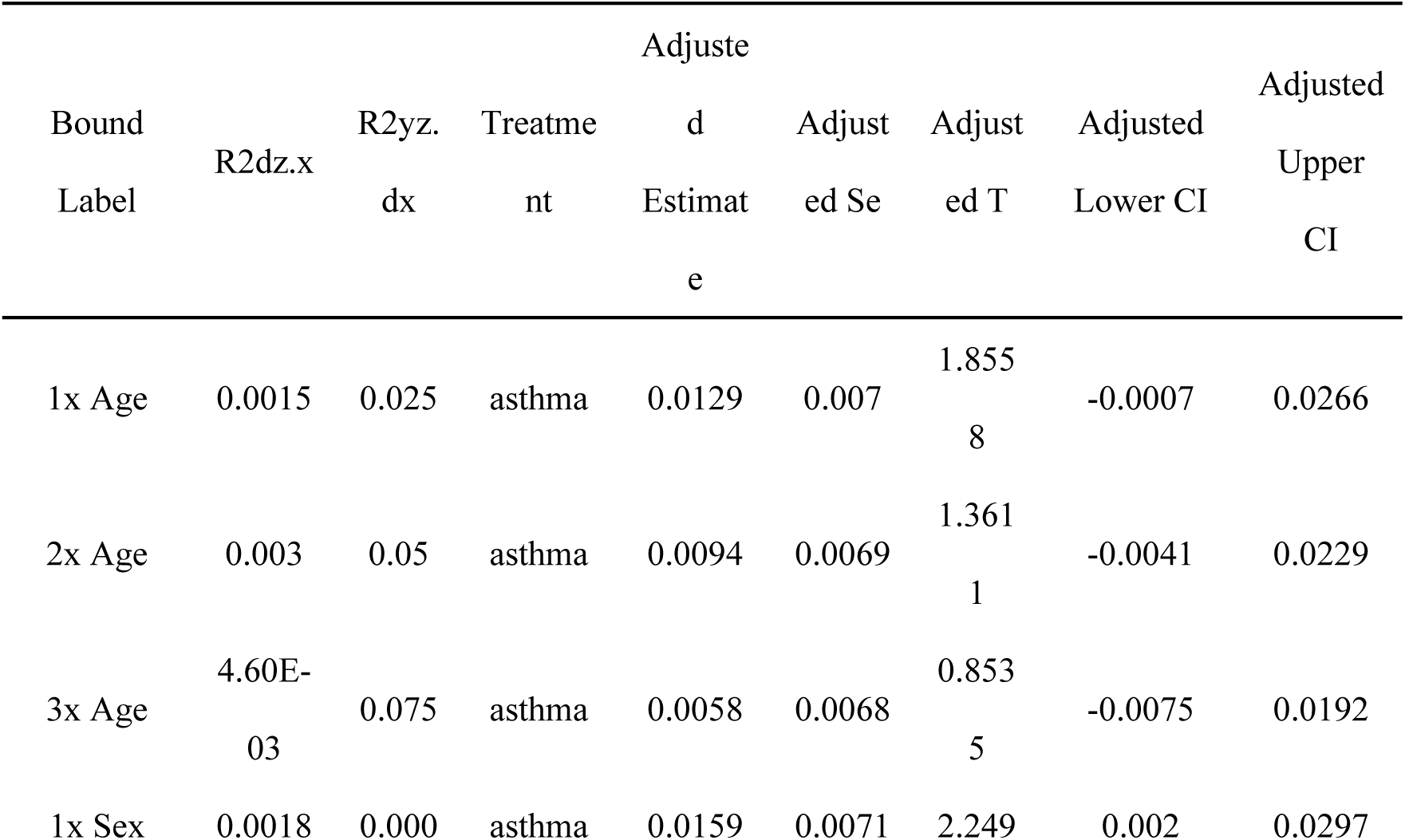
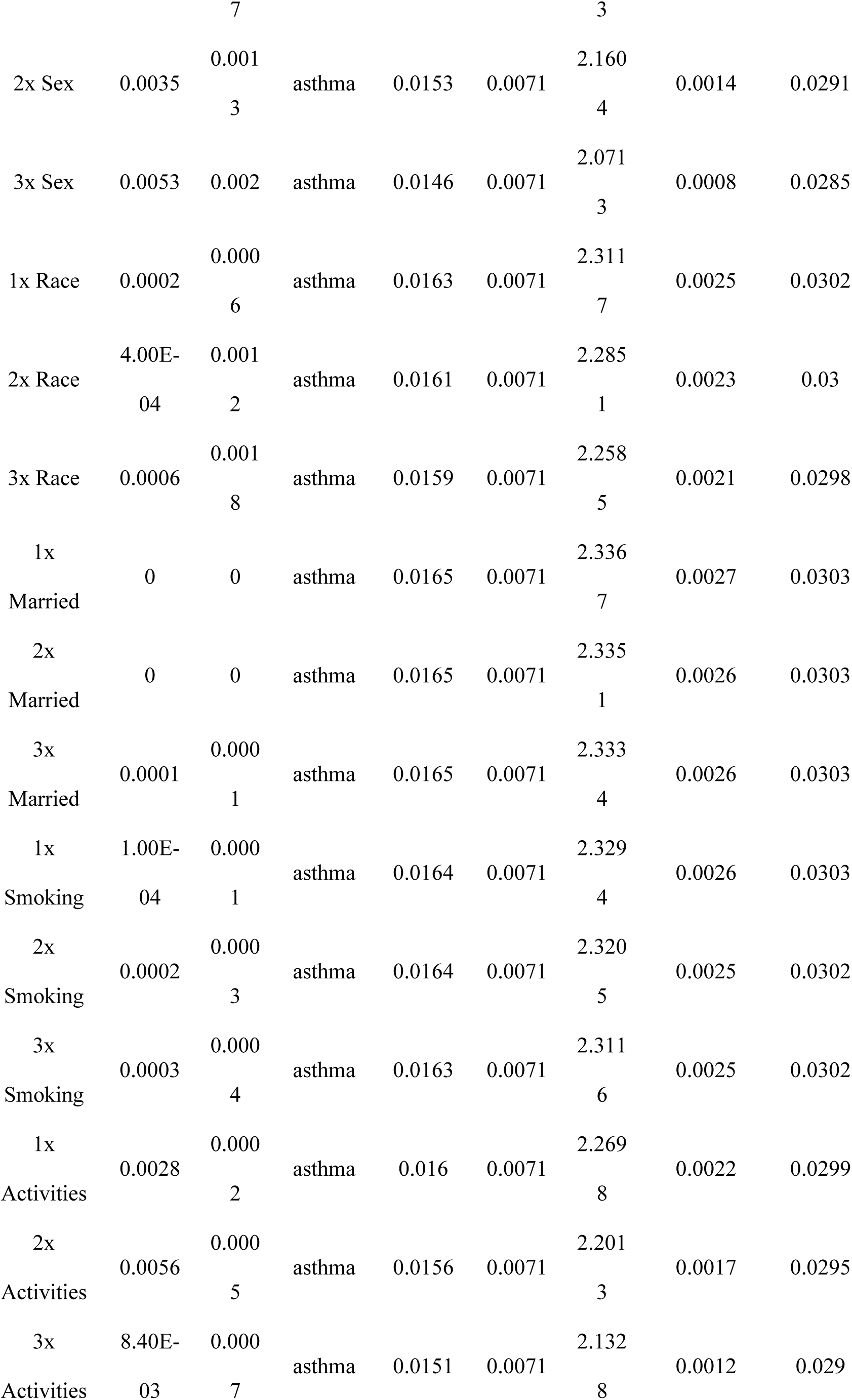
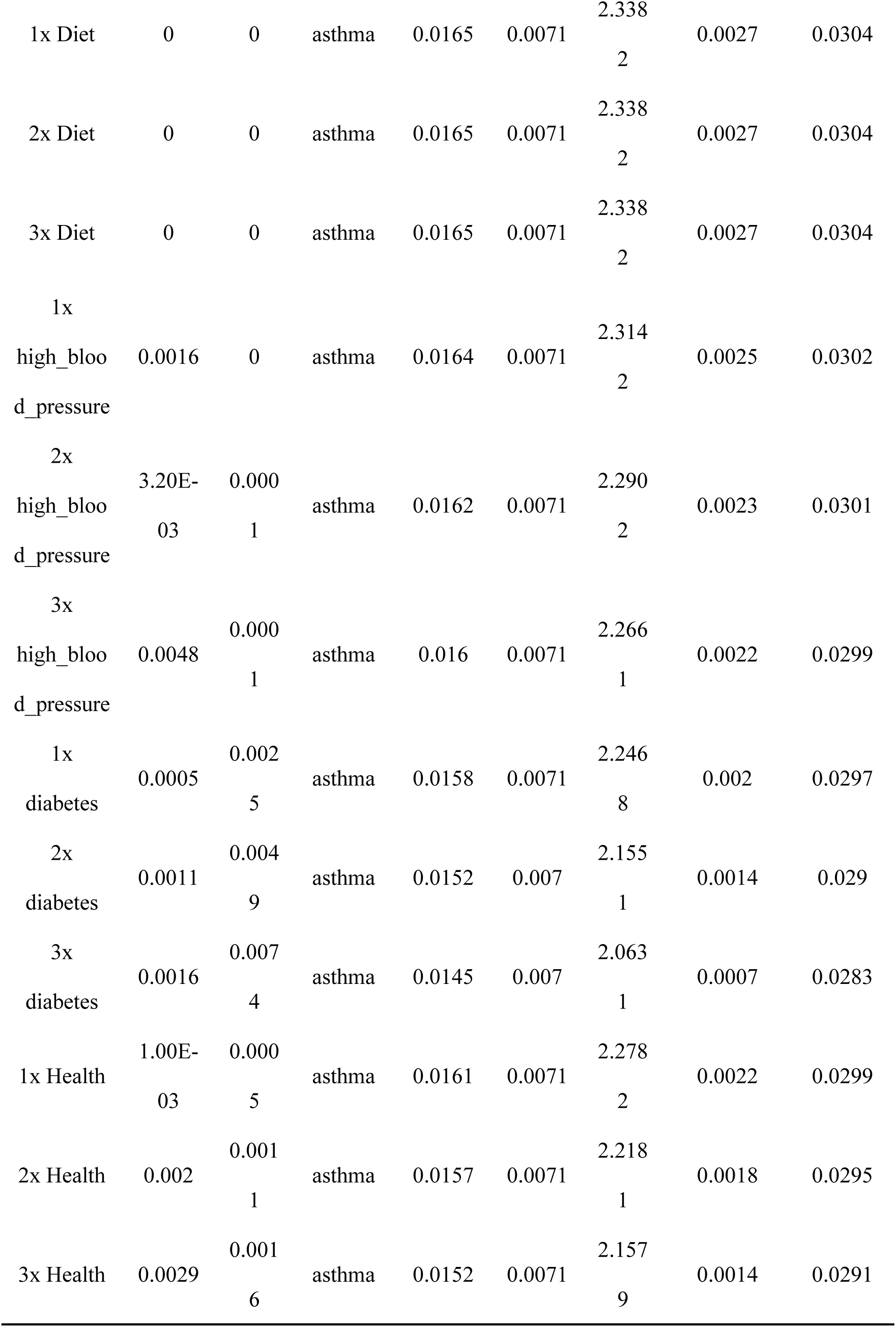
Univariate benchmark sensitivity analysis results: Bound Label: benchmark strength identifier. R²_{dz.x}: partial coefficient of determination between unobserved variable and treatment variable (asthma), representing the proportion of residual variance in the treatment variable explained by the unobserved variable after adjustment for observed variables. R²_{yz.dx}: partial coefficient of determination between unobserved variable and outcome variable (MD), representing the proportion of residual variance in the outcome variable explained by the unobserved variable after adjustment for observed variables and treatment variable. Treatment: name of treatment variable (asthma). Adjusted Estimate: adjusted coefficient estimate, reflecting the effect size of asthma on MD in the presence of unobserved confounding at a specified strength. Adjusted Se/T: adjusted standard error and t-value for statistical inference. Adjusted Lower/Upper CI: lower/upper bounds of the adjusted 95% confidence interval.

## 4. Discussion

In this study, asthma was significantly associated with MD across multiple analytic approaches. In adjusted logistic regression models, the association between asthma and MD remained positive after accounting for demographic, lifestyle, and health-related covariates. Subgroup analyses further suggested that this association was consistent across different demographic and clinical subgroups, although the magnitude of the association varied between subgroups, with a stronger association observed among participants aged over 70 years. In addition, ROC curve analysis indicated that asthma showed moderate discriminatory performance for MD in the fully adjusted model. Smooth curve analysis demonstrated a significant positive dose-response relationship across the observed range, suggesting asthma may serve as a key precipitating factor for MD. Sensitivity and benchmark analyses revealed that the observed association was robust to unmeasured confounding, although strong unobserved confounding such as age, sex, and health status could potentially influence the estimated effect. Taken together, our findings indicated that asthma remained positively associated with MD after adjustment for comprehensive covariates, suggesting that asthma itself or its underlying pathophysiological processes may serve as a key risk factor for MD.

The observed association between asthma and MD persisted after adjustment for a comprehensive set of covariates, including age, sex, ethnicity, marital status, smoking status, hypertension, diabetes, diet, physical activity, and health status (Model 3: OR > 1, p < 0.05). Our finding can be interpreted in the context of emerging evidence from international cohort studies examining the link between respiratory diseases and MD. For example, the Beaver Dam Eye Study revealed that after adjusting for age, sex, and smoking, pulmonary diseases and respiratory symptoms were independently associated with AMD progression—particularly for exudative (wet) MD (OR≈3.65)[32]. However, findings have not been entirely consistent across study designs and populations. A Taiwanese case-control study reported an overall association between allergic diseases and AMD but no statistically significant correlation for asthma alone (aOR=0.99, 95% CI: 0.93–1.06)[18]. In contrast, a recent nationwide cohort from Korea observed elevated risks of allergic diseases, including asthma, in AMD patients[19]. Overall, existing studies support a possible association between asthma and MD, although findings across studies and populations have not been entirely consistent.

In the present analysis, several established risk factors for MD showed associations consistent with prior reports, supporting the internal coherence of our findings[4, 32]. Age showed a strong, graded association with MD, in line with previous studies demonstrating that AMD prevalence increases sharply after 50 years, with late-stage AMD particularly common in those over 70[5, 33, 34]. Smoking, hypertension, diabetes, and poorer self-rated health were all more prevalent among participants with MD and remained associated with MD risk in multivariable models, consistent with existing AMD literatures. Even after adjusting for all these known covariates, the asthma-MD association remained statistically significant and robust across multiple models. This observation raises the possibility that asthma itself, or its underlying pathophysiological processes may contribute, directly or indirectly, to the development of MD.

Several biological mechanisms may help explain the observed association between asthma and MD. Asthma stands as a chronic inflammatory condition marked by systemic enrichment of inflammatory mediators—including interleukin-6, tumor necrosis factor-α, and C-reactive protein—all of which have been linked to endothelial dysfunction and microvascular damage in other organ systems[35]. Increasing evidence indicates that AMD also involves chronic inflammatory and immune-mediated processes, including complement activation, para-inflammation at the retinal pigment epithelium (RPE)-choroid interface, and microglial dysregulation[36–38]. In this context, systemic inflammation and immune dysregulation in asthma may amplify local inflammatory processes in the macula, accelerating drusen formation, RPE dysfunction, and progression to clinically manifest MD in susceptible individuals[39, 40].

Beyond inflammation, oxidative stress and hypoxia may represent additional pathways linking asthma with MD. Patients with asthma often experience intermittent hypoxemia during exacerbations, along with chronic oxidative stress driven by airway inflammation and increased production of reactive oxygen species. These systemic alterations can impair mitochondrial function, reduce antioxidant capacity, and promote lipid peroxidation in RPE cells and photoreceptors[41–44]—all processes have been identified as key contributors to AMD pathophysiology.

Previous clinical and experimental studies indicates that hypoxia is associated with alteration in posterior ocular hemodynamics and oxygenation, disruption of choroidal-retinal perfusion homeostasis, upregulation of vascular endothelial growth factor, and an increased tendency of neovascularization risk, particularly in the context of aging[45]. In our study, the association between asthma and MD appeared stronger among older participants. One possible explanation is that age-related declines in choroidal perfusion reserve and Bruch’s membrane integrity, may increase macular susceptibility to the downstream effects of systemic hypoxia and oxidative stress.

In addition to these biological mechanisms, methodological considerations such as indication confounding and drug effects warrant attention. Systemic and inhaled corticosteroids are widely used to manage asthma, and long-term exposure is a well-established risk factor for ocular hypertension, cataracts, and central serous chorioretinopathy. However, existing observational studies have no consistent evidence demonstrating a substantial effect of corticosteroids on MD risk after adjustment for baseline underlying disease severity[46, 47]. In this analysis, we adjusted for major comorbidities and lifestyle factors, and sensitivity analyses suggested that relatively strong unmeasured confounding would be necessary to fully attenuate the observed association. Nevertheless, potential confounding effect of corticosteroid use cannot be entirely excluded. Studies with more detailed data on asthma severity, acute exacerbation frequency, pulmonary function, and corticosteroid use may help clarify the independent contributions of disease activity and treatment exposure.

This study was based on data from a large, nationally representative cohort with standardized data collection and comprehensive covariate information. Multiple analytical approaches were applied, including multivariate logistic regression, ROC curve analysis, stratified analyses, and sensitivity analyses, to assess the association between asthma and MD, strengthening the robustness and credibility of the results. However, several limitations should be considered. First, the analysis mainly focused on the older adult population in the UK, which may limit the generalizability of the findings to younger populations or to individuals from other geographic or ethnic backgrounds. Second, the available data lacked detailed information on asthma phenotypes, disease severity, and MD subtypes. As a result, this limitation hindered in-depth exploration of potential dose–response relationships and phenotype-specific associations. Third, owing to the observational design, causal inference cannot be established. We cannot exclude the possibility of reverse causation, residual confounding, or it is also possible that asthma and MD share common upstream determinants, rather than one directly causing the other.

Despite these constraints, our findings may have implications for clinical practice and public health. For older adults with asthma—particularly adults ≥70 years with hypertension, diabetes, or smoking history—clinicians should recognize their potentially increased MD risk. In clinical settings, more attention to macular-related symptoms—such as central visual acuity changes and reading difficulties—may facilitate early detection and intervention of MD for these populations. Future studies in other large, ethnically diverse cohorts, prospective cohort studies or randomized controlled trials, may help investigate whether optimized asthma control or targeted therapies can reduce the incidence or progression of MD. Studies combining systemic inflammatory and oxidative stress biomarkers, multimodal macular imaging, genetic profiling, and detailed medication records could provide novel insight into pathways linking asthma and macular health.

In conclusion, this study identified a consistent association between asthma and MD in older adults. These findings add to the existing literature on allergic respiratory diseases and MD, and highlight the relevance of considering ocular outcomes in the long-term management of patients with chronic asthma.

## Declarations

## Data Availability

The raw data and materials generated in this study have been deposited in the FigShare repository. The data are publicly accessible at: https://doi.org/10.6084/m9.figshare.31305859, where 10.6084/m9.figshare.31305859 is the permanent digital object identifier (DOI) assigned to this dataset.

## Acknowledgements

We would like to express our sincere gratitude to all individuals and organizations who supported and assisted us throughout this research. Special thanks to the following authors: In conclusion, we extend our thanks to everyone who has supported and assisted us along the way. Without your support, this research would not have been possible.

## Author Contributions

Yini Yang: Conceptualization, Methodology, Software, Validation,

Formal analysis,Investigation, Data Curation, Writing - Original Draft,Visualization

Jingwei Li: Resources, Supervision, Writing - Review & Editing, Funding acquisition.

All authors have read and agreed to the published version of the manuscript.

## Ethics Statement

This study was a retrospective analysis of publicly available data and did not require ethical approval.

## Availability of data and materials

All relevant data are available within the paper and its Supporting Information files. The raw data and materials generated in this study have been deposited in the FigShare repository. The data are publicly accessible at: https://doi.org/10.6084/m9.figshare.31305859, where 10.6084/m9.figshare.31305859 is the permanent digital object identifier (DOI) assigned to this dataset.

## Supporting information

**S1 Table.** Variables and coding information: All variables were extracted from the English Longitudinal Study of Ageing (ELSA) database, with variable definitions and codes corresponding to original variable codes in the database.

